# Effect of continuous anesthetic wound infusion on postoperative activity in patients who underwent midline laparotomy for gynecological diseases

**DOI:** 10.1101/2023.02.19.23286163

**Authors:** Min Kyung Kim, Kidong Kim, Youngmi Park, Dong Hoon Suh, Jae Hong No, Yong Beom Kim

## Abstract

**Objective:** This study aimed to assess whether continuous anesthetic wound infusion (CAWI) increases postoperative activity in patients who undergo midline laparotomy for gynecological diseases.

**Methods:** This was an unplanned secondary analysis of a randomized controlled trial examining the effect of an activity tracker with feedback on postoperative activity in patients who underwent midline laparotomy for gynecological diseases (NCT02025387). Of the 53 patients included in the trial (12 patients used CAWI and 41 patients did not), 11 patients with CAWI (case group) and 11 without CAWI (control group) were selected by matching the duration of surgery. We compared the postoperative activity and other endpoints between the case and control groups.

**Results:** The case and control groups had similar baseline and surgical characteristics. The recovery rate on postoperative day 4 (step counts at day 4/ daily step counts at preoperative baseline) of the case group was numerically higher, but not significantly different from that of the control group (58% vs. 44%; p=0.450). Least, average and present pain on postoperative day 2 were lower in the case group than in the control group. Pain on postoperative day 5, fatigue, gas out, soft blend diet initiation, ileus, and length of hospital stay were similar between the case and control groups.

**Conclusions:** In this exploratory analysis, CAWI was not associated with increased postoperative activity in patients who underwent midline laparotomy for gynecological diseases.

## Introduction

Postoperative activity is associated with complications and recovery. For example, one systemic review showed that the amount of postoperative activity is negatively correlated with the length of hospital stay [1]. Bed rest after surgery increases the chances of pulmonary embolism and deep vein thrombosis [2, 3]

It has been suggested that increasing postoperative activity is beneficial. For example, early ambulation after open abdominal surgery prevents postoperative complications and reduces the length of stay [1]. In patients who started walking within 3 days after surgery, the time to independent standing and walking was significantly shortened [4]. Early ambulation also prevents venous thromboembolism [5-7].

Postoperative pain is one of the main hurdles in increasing postoperative activity. A negative correlation between pain and activity after cesarean section was previously reported [8]. Another study reported that postoperative daytime steps increased as postoperative pain was relieved [9]. Similarly, postoperative pain relief in hip fracture patients was shown to promote early functional gait recovery [10]. Such a positive correlation was also seen in patients undergoing knee arthroplasty, where postoperative pain management was associated with better range of motion, better performance in daily activities, and faster gait speed [11].

Continuous anesthetic wound infusion (CAWI) is a device that continuously releases a local anesthetic into the surgical site. The CAWI has been reported to effectively relieve postoperative pain. According to a meta-analysis that included 121 randomized controlled trials, CAWI was reported to reduce pain during laparotomy and sternotomy [12]. In another study, CAWI reduced additive opioid consumption [13]. We also reported that CAWI reduced pain in patients with gynecologic cancer [14].

We hypothesized that CAWI might increase postoperative activity through pain relief. This study therefore aimed to assess whether CAWI increases postoperative activity in patients who underwent midline laparotomy for gynecological diseases.

## Materials and methods

### Study information and population

This is an unplanned secondary analysis of a randomized controlled trial examining the effect of an activity tracker with feedback on postoperative activity in patients who underwent midline laparotomy for gynecological diseases [15]. The recovery rate of activity was measured by comparing the number of steps taken before and after midline laparotomy. The secondary analysis was approved, and the requirement for informed consent was waived by the Institutional Review Board of our institute (IRB No. B-2108-700-106).

The medical records of the 53 patients who participated in the randomized controlled trial were reviewed. Twelve patients received both CAWI and intravenous patient-controlled analgesia (PCA), but 41 patients received only intravenous PCA. By considering the duration of surgery as a factor to match patients receiving CAWI with those receiving PCA alone (nearest matching duration within ± 30 min), 11 patients with CAWI (case group) and 11 patients without CAWI (control group) were selected (Figure 1).

**Figure 1.**
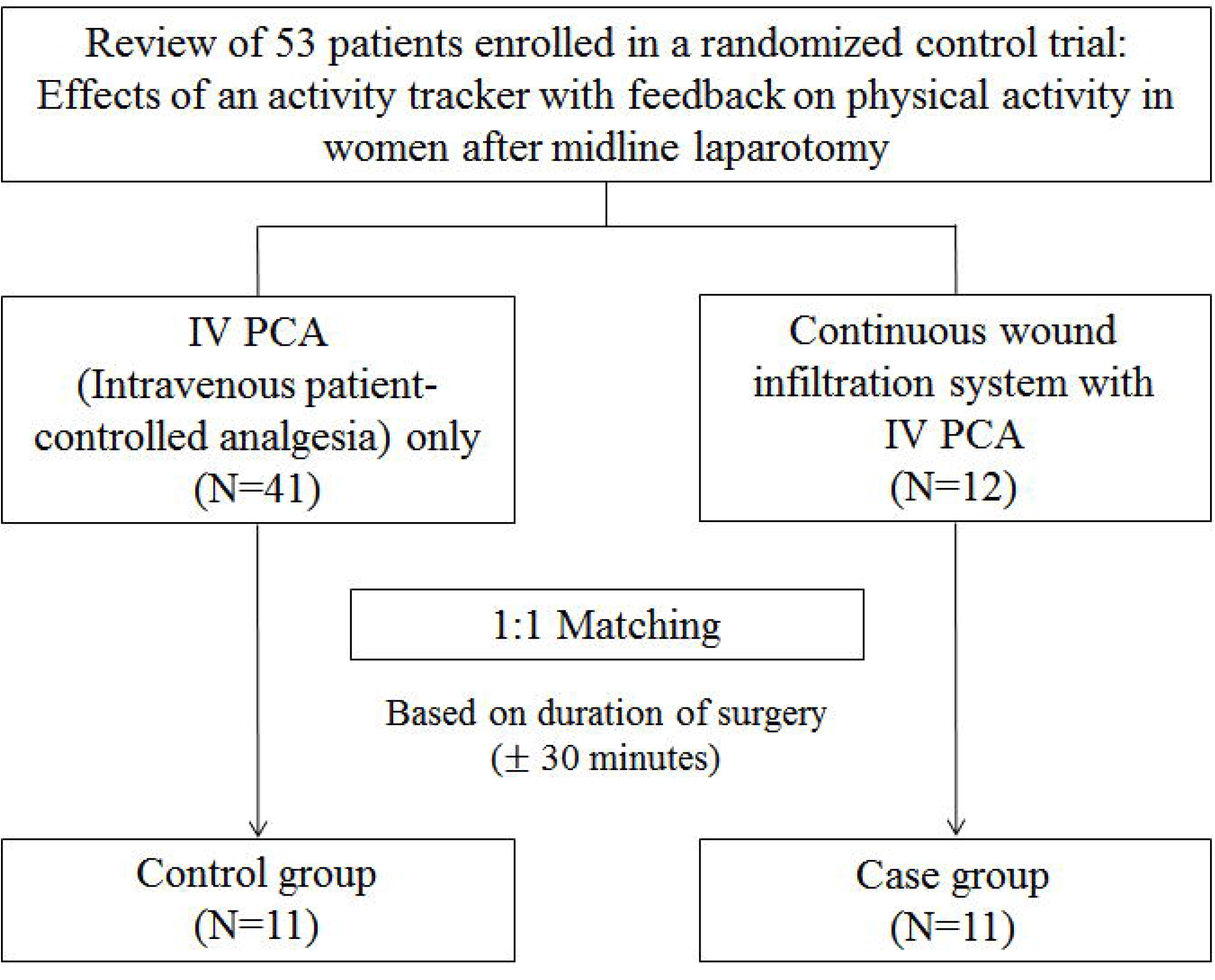
Schematic flow of patient selection criteria

### Objectives

The primary objective was to compare the recovery rate on postoperative day 4 (step counts on postoperative day 4 divided by the daily step counts at preoperative baseline) between groups. The secondary objectives were to compare the recovery rate over time, pain measured using the Brief Pain Index (BPI), fatigue measured with a brief fatigue index (BFI), gas out day, soft blend diet initiation, ileus rate, and length of stay.

### Statistical methods

The Mann-Whitney test or independent t-test was used to compare continuous variables. Chi-square tests were used to compare the categorical variables. After conversion to the log scale, a linear mixed model was used to analyze the recovery rate over time. *P*-value of < 0.05 was considered statistically significant. All analyses were performed using SPSS 25.0 (IBM, Armonk, NY, USA).

## Results

### Patient characteristics

The patient characteristics were similar between the case and control groups (Table 1). Specifically, the mean age was in the mid-50s in both the case and control groups. Height and weight of the case group were similar to those of the control group. The distribution of the American Society of Anesthesiologists (ASA) score was also similar between the groups. The duration of surgery was > 3 h, and the incision length was > 20 cm in both groups. The distribution of the arms in the randomized controlled trial was similar between the case and control groups.

**Table 1.** Patient characteristics

|  | Case<br>(N = 11) | Control<br>(N = 11) | p value |
| --- | --- | --- | --- |
| Age, mean ± SD (years) | 53.5 ± 11.1 | 57.5 ± 9.5 | 0.823 |
| Height, mean ± SD (cm) | 153.7 ± 6.4 | 157.7 ± 5.9 | 0.492 |
| Weight, mean ± SD (kg) | 52.8 ± 8.2 | 59.8 ± 7.3 | 0.207 |
| ASA score, n (%) |  |  | 0.392 |
| 1 | 6 (55) | 4 (36) |  |
| 2 | 5 (45) | 7 (64) |  |
| Duration of surgery, median (range) (min) | 235 (190-626) | 205 (162-615) | 0.562 |
| *Incision length, mean ± SD (cm) | 23.0 ± 6.4 | 20.6 ± 4.3 | 0.517 |
| Arm in randomized controlled trial, n (%) |  |  | 0.665 |
| Experimental arm | 6 (55) | 7 (64) |  |
| Control arm | 5 (45) | 4 (36) |  |

\*Based on 18 patients due to missing data. ASA, American Society of Anesthesiologists; SD, standard deviation.

### Postoperative outcomes

A comparison of the postoperative outcomes is summarized in Table 2. Step counts at the preoperative baseline and postoperative day 4 were similar between groups. The recovery rate on postoperative day 4 in the case group was numerically higher but not significantly different from that of the control group (58% vs. 44%; p=0.450).

**Table 2.** Postoperative outcomes

|  | Case<br>(N = 11) | Control<br>(N = 11) | p value |
| --- | --- | --- | --- |
| Daily step counts |  |  |  |
| Baseline, median (range) | 4053<br>(1203-18548) | 6209<br>(868-17516) | 0.670 |
| Postoperative day 4, mean ± SD | 2864±2074 | 2389±1916 | 0.583 |
| Recovery rate at postoperative day 4, median (range) (%) | 58 (3.5-201.1) | 44 (0.4-247.5) | 0.450 |
| BPI at postoperative day 2 |  |  |  |
| Worst in the last 24 h, mean ± SD | 7.1±2.0 | 8.1±1.7 | 0.217 |
| Least in the last 24 h, median (range) | 2 (0-7) | 4 (2-9) | 0.016 |
| Average, median (range) | 4 (2-8) | 5 (4-10) | 0.047 |
| Present, mean ± SD | 4.1±2.3 | 6.3±2.1 | 0.029 |
| BPI at postoperative day 5 |  |  |  |
| Worst in the last 24 h, median (range) | 3 (0-6) | 7 (3-10) | 0.813 |
| Least in the last 24 h, median (range) | 5 (4-8) | 2 (1-5) | 0.681 |
| Average, mean ± SD | 4.9±1.4 | 4.8±2.0 | 0.904 |
| Present, mean ± SD | 4.0±1.8 | 4.5±1.8 | 0.638 |
| BFI at postoperative day 5 |  |  |  |
| Present, median (range) | 5 (5-7) | 5 (3-9) | 0.754 |
| Usual during past 24 h, median (range) | 5 (4-8) | 5 (3-8) | 0.151 |
| Worst during past 24 h, mean ± SD | 7.5±1.9 | 6.8±2.3 | 0.491 |
| Gas out day, mean ± SD | 3.3±1.1 | 3.1±0.8 | 0.426 |
| Soft blend diet initiation, median (range) | 2 (1-4) | 1 (1-5) | 0.680 |
| Ileus, n (%) | 1/11 (9) | 0/11(0) | 0.306 |
| Length of stay, (days), mean ± SD | 14.0±15.2 | 12.0±4.6 | 0.418 |

BPI, Brief Pain Inventory; BFI, Brief Fatigue Inventory; SD, standard deviation

Least, average, and present pain measured with BPI on postoperative day 2 was lower in the case group than in the control group. However, the BPI scores on postoperative day 5 were similar between groups. Fatigue, gas out days, soft blend diet initiation days, ileus, and length of stay were all similar between groups.

### Recovery rate over time

The recovery rate over the postoperative day showed a gradual increase in the case group, but a fluctuation in the control group (Figure 2). Linear predictions of the log-transformed recovery rate over the postoperative day were also similar between the case and control groups (p = 0.584, Figure 3).

**Figure 2.**
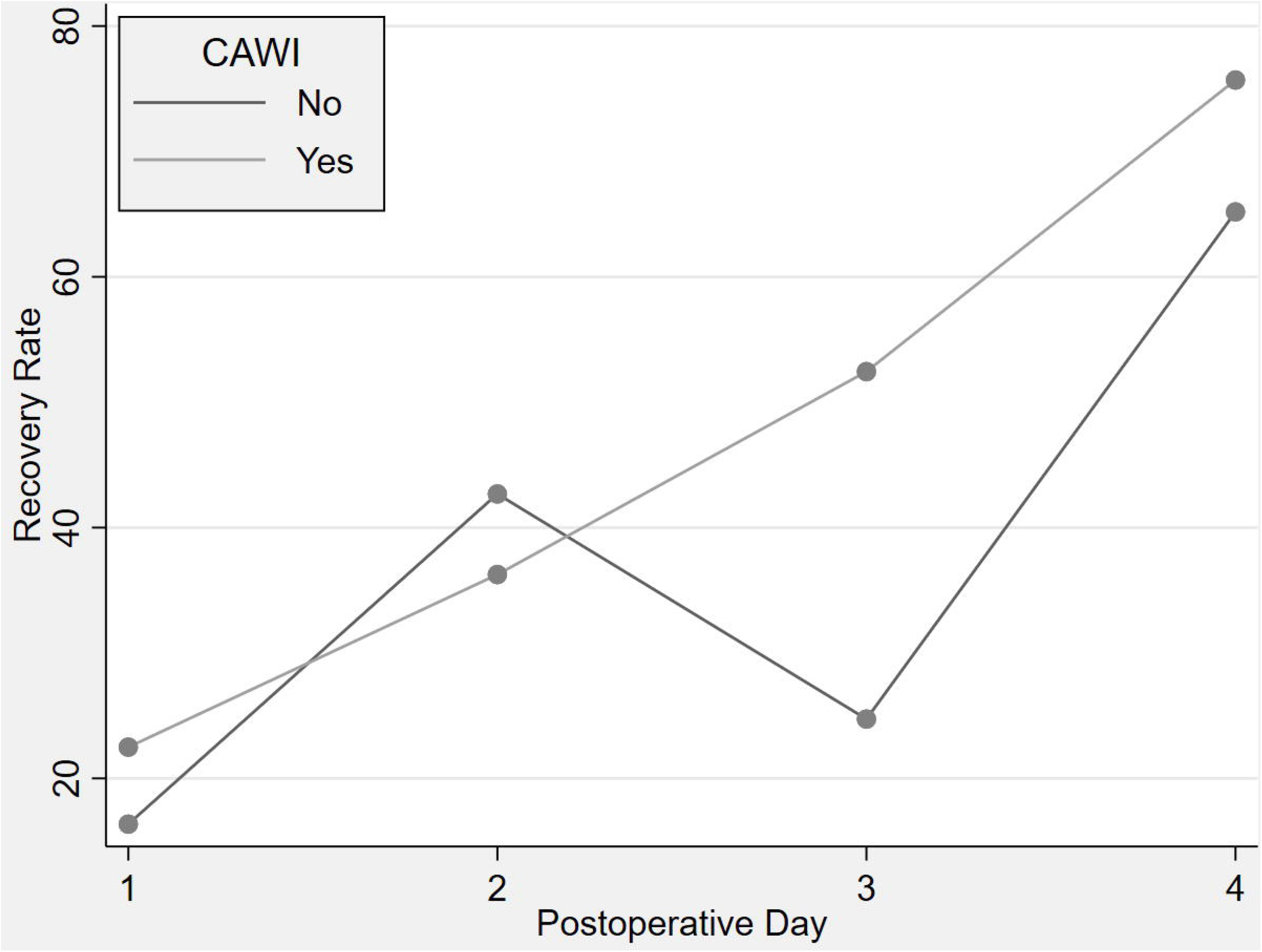
Recovery rate over postoperative days The recovery rate over the postoperative day showed a gradual increase in the case group, but a fluctuation in the control group.

**Figure 3.**
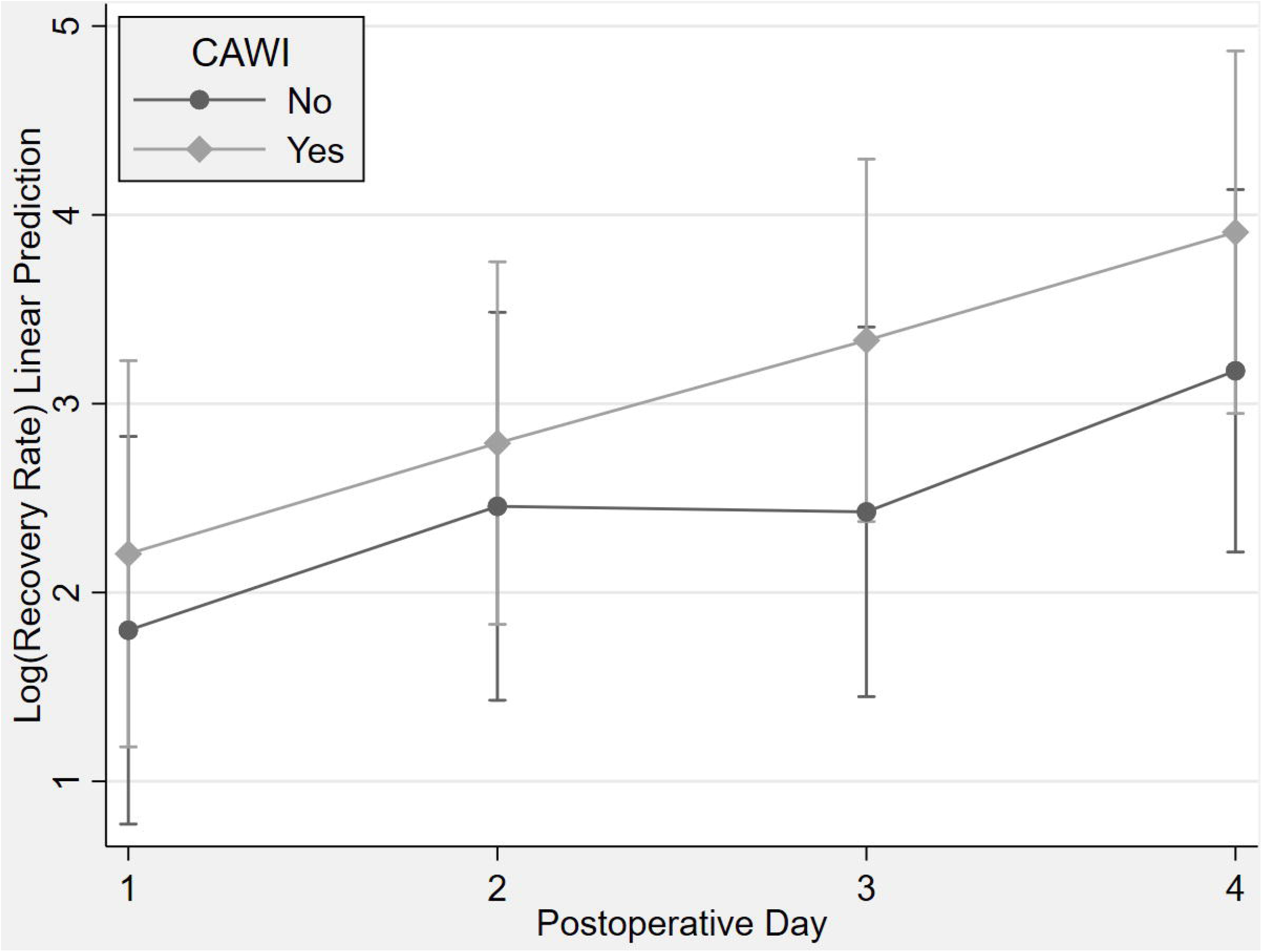
Linear prediction of log-transformed recovery rate Linear predictions of the log-transformed recovery rate over the postoperative day were similar between the case and control groups.

## Discussion

### Main findings of our study

A previous study reported that one of the major reasons for a quick postoperative recovery is controlling acute postoperative pain [11]. In one study, postoperative activity increased, whereas the pain decreased over time, after laparotomy surgery. Therefore, we hypothesized that CAWI might increase postoperative activity through pain relief. However, in this study, the CAWI was not associated with increased postoperative activity. Although not statistically significant, the recovery rate on postoperative day 4 was higher in the case group than in the control group. The small sample size could be a reason for the negative results.

### CAWI reduces pain

In this study, CAWI was associated with reduced pain on postoperative day 2, but not on day 5. Similar results have been reported in previous studies. For example, a study reported that acute-phase pain control was effective in the CAWI group, especially for laparotomy surgery [12]. Previous studies also have shown earlier retention of bowel function and a shorter hospital stay with CAWI [1, 13], but there were no significant differences observed in our study.

### Advantages of our study

The strength of our study was that the amount of postoperative activity and degree of postoperative pain could be quantitatively assessed using an objective number. There have been only a few studies investigating the relationship between postoperative pain and activity. In our study, activity accuracy was assessed using an activity tracker and pain through the BPI score. The basal activity was quantified using an activity tracker to assess the recovery rate.

### Limitations of our study

A limitation of our study was the possibility of selection bias. Specifically, the CAWI tended to be applied to longer and major surgeries, which led to differences in characteristics between the two groups. We attempted to negate the selection bias using duration of surgery as a factor for matching. In addition, the number of patients included in our study was insufficient to achieve significant results. In future studies, enrolling higher number of participants may yield significant results.

### Conclusion

In this exploratory analysis, CAWI was not associated with increased postoperative activity in patients who underwent midline laparotomy for gynecological diseases.

## Data Availability

All data produced in the present study are available upon reasonable request to the authors

## Acknowledgments

The authors thank the Division of Statistics in the Medical Research Collaborating Center at Seoul National University Bundang Hospital for the statistical analysis. We would like to thank Editage (www.editage.co.kr) for English language editing.

## Ethical approval

This study was performed after obtaining approval of the Institutional Review Board of Seoul National University Bundang Hospital (IRB No. B-2108-700-106). This study was performed in accordance with the principles of the Declaration of Helsinki.

## Patient consent

Written informed consent and the use of images from the patients were not required for publication.

